# SARS-CoV-2 infection risk in COVISHIELD (AstraZeneca-SII) Vaccinated Healthcare Workers in a Tertiary Cancer Care Centre of North-East India

**DOI:** 10.1101/2023.12.11.23299808

**Authors:** Dhruba Jyoti Sarma, Biki Chandra Das, Avdhesh Kumar Rai, Pallavi Sarma, Priyanki Baruah, Kamalika Bhandari Deka, Mouchumee Bhattacharyya, Anupam Sarma, Debanjana Barman, Amal Chandra Kataki, Sawmik Das, Rashmisnata Barman

## Abstract

COVID-19 vaccination of Healthcare workers and vulnerable groups remain priority in containing the spread of SARS-CoV-2 infection and health complications arising post COVID-19. In India, COVISHIELD is the major part of COVID-19 vaccination drive. post-vaccination SARS-CoV-2 infection risk and its impact on severity as well as mortality need to be assessed in vaccinated population. The study included 350 COVISHIELD vaccinated HCWs in tertiary cancer care centre of North-East India. A total of 16 symptomatic HCWs (4.57%) were confirmed as SARS-CoV-2 positive. Among SARS-CoV-2 positive FV HCWs none were positive within 14 days, 13.33 % within 15 to 30 days, 53.33% within 30 to 45 days, and 33.33% after 45 or more days. None of the SARS-CoV-2 positive HCWs required oxygen support or hospitalisation. We also found that 66.67% FV HCWs (n=10) had Ct values below 20, 20% (n=3) had Ct values in between 20-25 and 13.33% (n=2) had Ct values in between 30 to 35. In North-East India COVISHIELD FV HCWs HCW,SARS-CoV-2 infection and COVID-19 symptoms were mild. Majority of FV HCWs have RT-PCR COVID-19 positive Ct Value was below 20. SARS-CoV-2 infection occurred mostly after 15 days to 45 days or more post vaccination.

## Introduction

India is currently facing the second wave of SARS-CoV-2 pandemic, even though the daily positivity rate is declining, but there are predictions for a possibility of a third wave. ^1^ India launched its first phase vaccination drive for the healthcare workers (HCWs) on 16th January 2021. Primarily the vaccination was started with COVISHEILD (Serum Institute India-AstraZeneca) and later, COVAXIN (Bharat biotech) was included in the vaccination drive. In May 2021, Sputnik V has also been approved by Government of India for emergency use authorisation (EUA). As per the current vaccination data up until 12^th^ June 2021, 246 million individuals have been vaccinated with at least one dose, wherein around 46.5 million (3.4%) are fully vaccinated (FV) with both doses (recipients of both doses of vaccination are designated as fully vaccinated (FV)). In the state of Assam, Northeast India; around 850,474 people (∼2.7% of the population) have been FV. There are various reports from many countries of the world that even after being FV, people have been infected with SARS-CoV-2 ^2-4^. However, the severity of the symptoms and hospitalisation requirements were low. Multiple variants of SARS-CoV-2 have emerged and many of these were reported in SARS-CoV-2 patients in India and around the world. Recently, WHO (World Health Organisation) has classified variants of concern (VOC) as Alpha, Beta, Gama and Delta. The huge surge of SARS-CoV-2 infections and mortality in the second wave is suggested to be driven by these VOC, especially delta variant. The VOCs of SARS-CoV-2 have higher transmission capability and may be able to evade the vaccine-induced immunity even in the FV individuals (11). Therefore, Postvaccination SARS-CoV-2 infection risk and its impact on severity as well as mortality need to be assessed in vaccinated population. Our institution, Dr B Borooah Cancer Institute (Dr BBCI) Guwahati; is a comprehensive tertiary cancer centre from Assam, Northeast India. All cancer patients have to undergo the RT-PCR test for SARS-CoV-2 before admission. In our hospital, all the Government of India SARS-CoV-2 guidelines are followed such as wearing a double mask; maintaining social distance, hand hygiene etc. Any sign of SARS-CoV-2 related symptoms in HCWs is promptly investigated by RT-PCR testing and if it is positive, treated for SARS-CoV-2.

## MATERIALS and METHODS

The study included 350 vaccinated HCWs of Dr BBCI. The included staff were categorised as Doctors, Nursing staff, Technical and Scientific staff, Administrative staff, Sanitation staff and Ward Attendants. The vaccination drive at Dr BBCI started with COVISHEILD on 12^th^ February 2021 and continued up to 4^th^ May 2021 in phases. From April 2021 to May 2021, SARS-CoV-2 infections were confirmed in post vaccinated symptomatic HCWs by RT-PCR using CoviPath COVID-19 RT-PCR Kit (Thermo Scientific, USA) as per manufacturer’s instruction. Viral RNA extractions from samples were done using QIAamp Viral RNA kit as per manufacturer’s instruction. Samples with Ct Value ≤35 for N gene, Orf1ab and RnaseP were considered positive.

## RESULTS

As summarised in table 1, out of 350 vaccinated HCWs (male 207, female 143), 330 (94.28%) were FV with COVISHIELD. A total of 16 symptomatic HCWs (4.57%) were confirmed as SARS-CoV-2 positive. Among SARS-CoV-2 positive HCWs, 93.75% (n=15) were FV and 6.25% (n=1) was vaccinated with a single dose. Among the SARS-CoV-2 positive HCWs fever was the primary reported symptom. However, none of the SARS-CoV-2 positive HCWs required oxygen support or hospitalisation.

**Table 1:** Summary of the SARS-CoV-2 Vaccination and SARS-CoV-2 Infection in HealthCare Workers (HCWs)

|  |  |  |  |  |
| --- | --- | --- | --- | --- |
| Vaccinated (at least one dose) |  | 350 |  |  |
| Fully vaccinated (both doses) |  | 330 (94.28% among vaccinated staff) |  |  |
| Total no of infection after vaccination (at least one dose) |  | 16 out of 350 (4.57% amongst vaccinated with at least one dose) |  |  |
|  |  | Male |  | 11 (68.75 %) |
|  |  | Female |  | 5 (31.25 %) |
| Viral load of infections for all 16 infections |  |  |  |  |
| Viral load | Ct value below 20 | Ct value 20-25 | Ct value between 25-30 | Ct value between 30-35 |
| No of infections | 11(68.75%) | 3 (18.75%) | 0 | 2 (12.5%) |
| Number of days after vaccination for all 16 infections |  |  |  |  |
|  | Within 0-14 days | Within 15-30 days | Within 30-45 days | More than 45 days |
| No of infections | 0 | 2 (12.5%) | 9 (56.25%) | 5 (31.25%) |
| Total no of infection after vaccination (both doses) |  | 15 out of 330 (4.54% amongst fully vaccinated) |  |  |
|  |  | Male |  | 11 (73.33 %) |
|  |  | Female |  | 4 (26.66 %) |
| Viral load of infections after both vaccination dose (15 infections) |  |  |  |  |
| Viral load | Ct value below 20 | Ct value 20-25 | Ct value between 25-30 | Ct value between 30-35 |
| No of infections | 10 (66.67%) | 3 (20%) | 0 | 2 (13.33%) |
| Number of days after vaccination after both vaccination (15 infections) |  |  |  |  |

|  | Within 0-14 days | Within 15-30 days | Within 30-45 days | More than 45 days |
| --- | --- | --- | --- | --- |
| No of infections | 0 | 2 (13.33%) | 8 (53.33%) | 5 (33.33%) |

We observed that among SARS-CoV-2 positive FV HCWs none were tested positive within 14 days. Within 15 to 30 days only 13.33 % were tested positive, within 30 to 45 days 53.33%, and 33.33% were tested positive for SARS-CoV-2 after 45 or more days.

When RT-PCR Ct values were evaluated in SARS-CoV-2 positive HCWs, we found that 66.67% HCWs (n=10) had Ct values below 20, 20% (n=3) had Ct values in between 20-25 and 13.33% (n=2) had Ct values in between 30 to 35. The single-dose vaccinated HCW was tested positive for SARS-CoV-2 on the 30^th^ day and had a Ct value below 20. The age distribution showed that out of the 16 HCWs, 50% (n=8) were in the age group of 40-50 years, 25% (n=4) HCWs were in the 30-40 years age group, 12.5 % each (n=2) in the age groups of below 30 and 50-60 age.

HCWs category wise distribution regarding vaccination and SARS-CoV-2 infection are summarised in Table2. Among SARS-CoV-2 positive HCWs, 37.5% were doctors (n=6), 31.25% (n=5) were nursing stuff, 18.75% (n=3) were technical and scientific staff and 12.5% (n=2) were administrative staffs. No infection was observed for the group of cleaning and sanitation staffs and ward attendants.

**Table 2:** HCWs Profile with reference to Vaccination and SARS-CoV-2 Infection.

| Profile of HCWs | No of vaccinated HCWs (%) | Number of SARS-CoV-2 positive HCWs (%) |
| --- | --- | --- |
| Doctor | 85 (24.28) | 6 (37.5) |
| Nursing staff | 67 (19.14) | 5 (31.25) |
| Technical and Scientific staffs | 64 (18.28) | 3 (18.75) |
| Administrative staff | 101 (28.85) | 2 (12.5) |
| Sanitation staff | 6 (1.71) | 0 |
| Ward attendants | 27 (7.71) | 0 |
| Total | 350 (100) | 16 (100) |

## DISCUSSION

The vaccination for SARS-CoV-2 has started in December 2020, there are few published reports regarding the infection of SARS-CoV-2 Postvaccination. We found in our study SARS-CoV-2 positivity rate among COVISHEILD vaccinated HCWs was 4.57% (16 out of 350). Amit et al.^2^ from Israel have reported that Pfizer-BioNTech RNA SARS-CoV-2 vaccine recipient HCWs had SARS-CoV-2 positivity rate of 0.54% (22 out of 4081). However, this study was conducted after the first dose only. Cucunawangsih et al^3^ reported an incidence of infection rate of 1.25% (13 out of 1040 HCWs) after both doses of the vaccine for CoronaVac. However none of the reported positive cases were beyond 14 days. They had shown that 53.84% HCWs had Ct values below 20, 30.76% had Ct values between 20 to 25, and 7.69% in each in the range of 25 to 30 and 30 to 35. None of the HCWs required hospitalisation. Further, Keehner et al.^4^ reported positivity rate of 0.0013% (37 out of 28184) for FV HCWs for Moderna mRNA-1273 and Pfizer BioNTech RNA vaccines. Out of those 37 FV HCWs, 22 (59.45%) had tested positive for SARS-CoV-2 within 8 days, 8 (21.62%), within 8-14 days and 7 (19.91%) post 15 days.

We found only one report from India by Tyagi et al.^5^ who have reported a positivity rate of 16.82% (18 out of 107) among the FV (COVAXIN or COVISHIELD) HCWs.^5^ 83.33% (15 out of 18) of the SARS-CoV-2 of the infections were found more than 15 days Postvaccination. Lumley et al.^6^ in a UK study observed among 8285 HCWs recipient of the Pfizer-BioNTech vaccine (1407 two doses) and 2738 HCWs recipients of the Oxford-AstraZeneca vaccine (49 two doses) that single dose vaccinated HCWs have reduction of 67% symptomatic SARS-CoV-2 infection. HCWs who received two doses (Pfizer-BioNTech vaccine and Oxford-AstraZeneca vaccine) has 90% and 85% reduced incidence of PCR-positive SARS-CoV-2 infection with or without symptoms.^6^ Shah et al. in UK Cohort which included 109,074 HCWs and 148,366 household members of HCWs who received at least one dose of the BNT162b2 mRNA (Pfizer-BioNTech) or ChAdOx1 nCoV-19 (Oxford-AstraZeneca) vaccine, Postvaccination SARS-CoV-2 positive HCWs were 0.99% (n=1086) and household members of HCWs were 0.77% (n=1152). Further, 64 household members of healthcare workers and 19 HCWs has COVID-19 hospitalisations Postvaccination (first dose) respectively.^7^ Faria et al. in Brazil cohort of HCWs (n=22,402), vaccinated with CoronaVac, found 1.69% (n=380) HCWs were positive for SARS-CoV-2 infection after first dose.^8^ In another study of BioNTech/Pfizer COVID-19 vaccine in Treviso Province, Italy among 6,423 HCWs. They reported, 84% effectiveness in preventing SARS-CoV-2 infection within the time intervals of 14–21 days from the first dose and 95% effectiveness in preventing SARS-CoV-2 infection atleast 7 days from the second dose.^9^ Thompson et al.^10^ reported that 0.04 SARS-CoV-2 infections per 1,000 person-days in m-RNA vaccine fully immunized persons (≥14 days after second dose). They found that among partially immunized persons (≥14 days after first dose and before second dose), 0.19 SARS-CoV-2 infections per 1,000 person-days occured.^10^

HCWs are very important and integral part of all global efforts towards prevention and management of COVID-19 pandemic. HCWs have been designated as priority group for COVID-19 vaccination in ongoing vaccination drives of various countries across the world. But still vaccine uptake hesitancy has been observed within different categories of HCWs regarding safety and efficacy of COVID-19 vaccines in preventing SARS-CoV-2 infection and risk of serious adverse events Postvaccination. Real world setting data of COVID-19 vaccines for HCWs (COVISHIELD, COVAXIN etc) regarding effectiveness and risk of serious adverse events is limited and necessitates requirement of **more** data. Such data in HCWs will be of immense benefit in boosting the acceptance of vaccines among HCWs. Though limitations of our study maybe, small sample size and absence of co-morbidities data in HCWs.

We conclude that low SARS-CoV-2 infection and COVID-19 symptoms in COVISHIELD fully vaccinated HCWs were mild in nature and COVID-19 hospitalizations were not required in any of the SARS-CoV-2 positive HCWs of North-East India. SARS-CoV-2 infection in HCWs occurred after 15 days Postvaccination. In view of emergence and identification of multiple VOCs of SARS-CoV-2 in India, further multi-centric studies with large cohort size maybe required to assess SARS-CoV-2 infections risk, morbidity and mortality due to COVID-19 in FV HCWs.

## Data Availability

None, All data is available in the tables

